# A novel cell type negatively associated with secondary autoimmunity in alemtuzumab-treated patients is revealed through single-cell longitudinal analysis of clinical trial samples

**DOI:** 10.1101/2021.06.21.21258814

**Authors:** Richa Hanamsagar, Mathew Chamberlain, Srinivas Shankara, Robert Marcus, Darren P. Baker, Evis Havari, Katherine Klinger, Frank O. Nestle, Emanuele de Rinaldis, Virginia Savova

## Abstract

Alemtuzumab, a humanized anti-CD52 monoclonal antibody, is an approved treatment for relapsing forms of multiple sclerosis (RMS). While its efficacy has been demonstrated in clinical studies, its use is associated with unpredictable non-MS autoimmunity manifesting months or years after treatment. Approximately 40% of treated patients present with autoimmune thyroid events, 2% with platelet deficiency (immune thrombocytopenia; ITP)^1^, and 0.34% with autoimmune nephropathies^2^. The lack of predictive biomarkers necessitates careful monitoring in clinical practice with a Risk Management Plan or Risk Evaluation and Mitigation Strategy (RMP/REMS) in place for early detection of these autoimmune events. We carried out a longitudinal single-cell analysis of PBMCs in a small subset of alemtuzumab-treated patients from the phase 3 CARE-MS I (CAMMS323) study^3^ and identified a novel platelet lineage cell negatively associated with thyroid autoimmunity. This discovery raises the possibility that a shared underlying mechanism may contribute to the incidence of thyroid autoimmunity and ITP in alemtuzumab-treated patients.

**Highlights:**

- A novel platelet-lineage cell type present in peripheral blood is uncovered and characterized
- Depletion and/or transcriptomic changes in this cell type are associated with increased incidence of secondary autoimmunity in MS patients on Alemtuzumab.
- Retrospective single-cell analysis studies can uncover novel insights and lead to future biomarker development

## Main

To understand if patients presenting with thyroid events following alemtuzumab treatment differed with respect to immune cell type composition, we profiled cryopreserved PBMC samples from 32 patients enrolled in CARE-MS I who received two alemtuzumab courses 12 months apart as per the clinical trial protocol (**Fig. 1A**). Blood samples were collected and processed before treatment (T0) and 24 months after the first treatment (T24). Of these patients, 11 presented with secondary autoimmune thyroid events (sAI) within the first 4 years of treatment, 18 patients remained free of such events and had no detectable autoantibodies (non-sAI), and 3 additional patients presented with autoantibody findings but no clinical manifestations (LA) (**Supp. File 1**).

**Figure 1:**
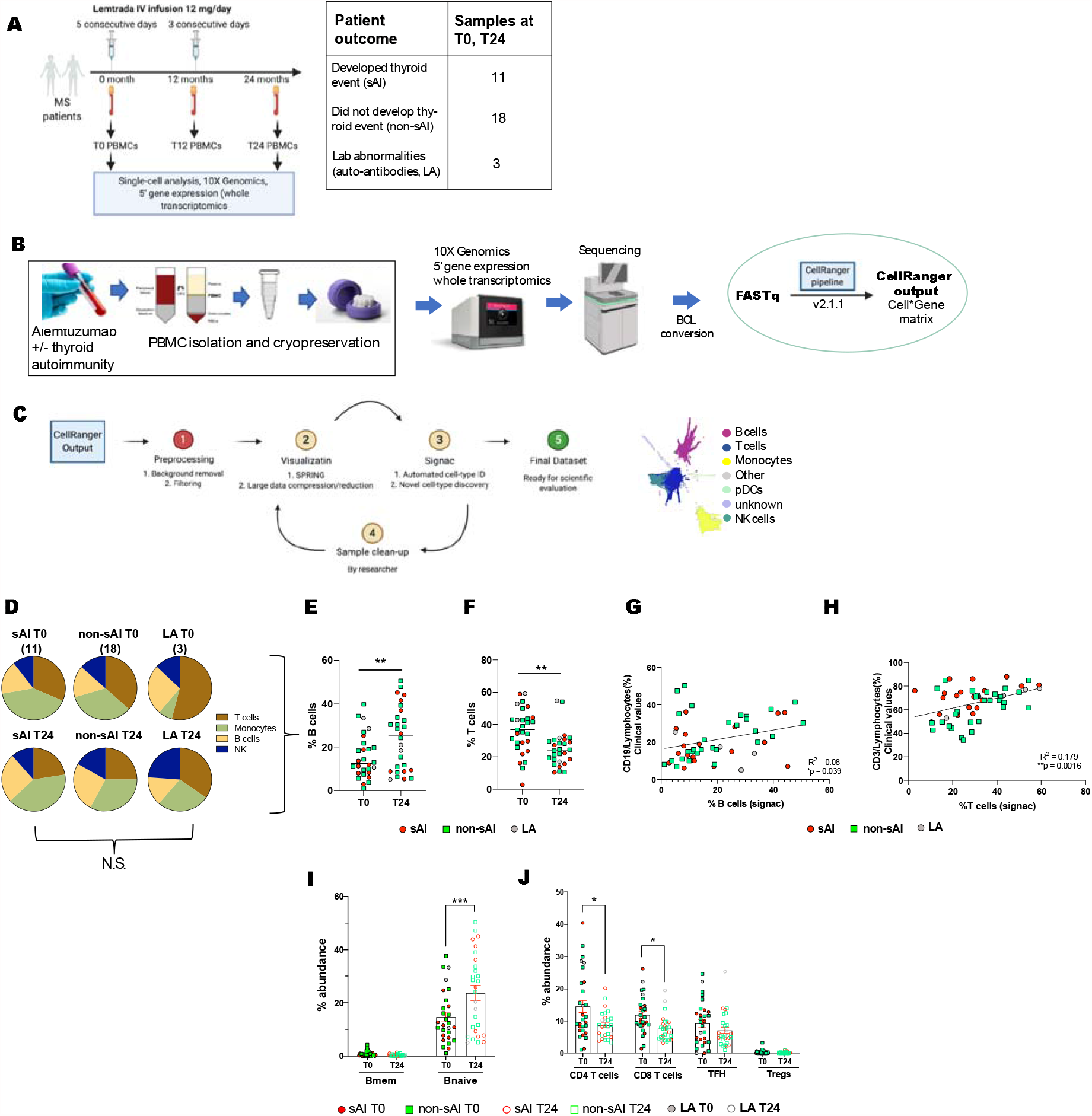
Preliminary single-cell transcriptomics analysis on whole PBMCs from alemtuzumab-treated MS patients. **A**. Single-cell transcriptomics was carried out on whole PBMC samples obtained from 32 MS patients at T0 and T24 post-treatment with alemtuzumab. Of these, 11 patients developed secondary autoimmunity in response to the treatment whereas 18 did not. Three patients displayed abnormal laboratory tests as indicated by the presence of auto-antibodies. **B**. Whole blood was collected from patients in CPT tubes containing Na-citrate, and PBMCs were isolated and cryopreserved. Cryopreserved PBMCs were thawed and counted as per 10X Genomics guidelines and run through the 10X Genomics Chromium device for encapsulation. Libraries were prepared using a 5’ gene expression reagent kit, and sequenced on the Novaseq sequencer. Following BCL conversion, FASTq files were processed through CellRanger (v2.1.1). **C**. The gene*barcode matrix files were preprocessed for removal of background RNA, empty barcodes, and mitochondrial genes using an internal pipeline. Dimensionality reduction and visualization were done with SPRING, and an automated cell-type annotation based on sorted bulk profiles was carried out; initially all samples were examined; bad quality samples with low cell type information content were eliminated from future analysis, and dimensionality reduction was re-applied to the remaining samples. The final dataset was used for all downstream scientific analysis. **D**. Pie charts depicting the distribution of main immune cell-types in single-cell data by Signac: T cells, Monocytes, B cells, and NK cells across the 6 disease groups: sAI T0 (secondary autoimmunity, time point 0, 11 samples), sAI T24 (secondary autoimmunity, time point 24 months), non-sAI T0 (no secondary autoimmunity, time point 0, 18 samples), non-sAI T24 (no secondary autoimmunity, time point 24 months), LA T0 (laboratory abnormalities, time point 0, 3 samples), and LA T24 (laboratory abnormalities, time point 24 months). **E, F**. Scatter bar plot depicting significant increase in % B cells and significant decrease in % T cells using single-cell data 24 months post first alemtuzumab treatment (Student’s t test, \**p* < 0.05 and \*\*\**p* < 0.001). **G, H**. Significant correlation between single-cell numbers and clinical values (flow cytometry) of % B cells (Pearson’s correlation R =0.08, \**p*<0.05) and % T cells (Pearson’s correlation R^2^ =0.179, *p*<0.0016) in all patients. **I, J**. Scatter bar plot depicting a significant increase in % of B naive cells and significant decrease in CD4 T- and CD8 T-cells in single-cell data post-alemtuzumab treatment (Mean±S.E.M; Two-way ANOVA with Sidak’s multiple comparison test \**p* < 0.05, \*\**p* < 0.001 and \*\*\**p* < 0.001).

The available samples were estimated and subsequently confirmed to contain a limited quantity of viable cells (2 × 10^2^-1 × 10^7^) which would not allow a comprehensive analysis of cell type composition using FACS or other techniques. We therefore carried out droplet-based parallel single-cell RNA sequencing (scRNA-seq) (**Fig. 1B**, **Supp. Fig. 1**). To process this dataset, we developed a high-throughput cloud-based pipeline with large dataset capacity to dimensionally reduce, visualize, and automatically label the data with known cell-type and subtype labels based on transcriptional similarity with sorted bulk profiles^4^ (**Fig. 1C**). We verified the automatically labeled annotations by confirming that marker expression on cell types and subtypes conformed to expectations (**Supp. Fig. 2**). To support the validity of the dataset, we examined changes in relative abundance of cell types before and after treatment (**Fig. 1D-F**). Consistent with previous observations^5^, we detected an increase in B-cells and a decrease in T-cells at the post-treatment timepoint which was 12 months after the second course. We were further able to correlate individual proportions of these cell-types in each sample to the clinically recorded lymphocyte count, showing that the scRNA-seq relative abundance measurement, as measured using the pipeline we developed, provided a reliable view on cell count, but offered a deeper resolution in terms of cell-types than standard FACS (**Fig. 1G-H**). No differences were observed in total monocyte and NK cell numbers in either group (**Supp. Fig. 3A-B**). NK cell abundance from single-cell data also strongly correlated with clinical values, further strengthening the possibility of using sc-data as a proxy for clinical values (**Supp. Fig. 3C**). Next, we compared B- and T-cell subtype differences in patients with and without thyroid events (sAI vs. non-sAI) before and after alemtuzumab treatment. Consistent with previous findings^5,6^, we found that numbers of naïve B-cells increased significantly, whereas CD4 and CD8 T-cell numbers reduced post treatment (**Fig. 1I-J**). No differences were observed in B- and T-cell subtypes in sAI versus non-sAI patients (**Supp. Fig. 4A-B**).

Since we observed no difference in the relative abundance of known cell types or subtypes between those with or without thyroid events, we analyzed the abundance of cell types classified as unknown by our automated algorithm. Surprisingly, this revealed a rare platelet-like cell-type (PLC) significantly lower in those with thyroid events compared to those without (**Fig. 2A-B**, sAI 0.07%; non-sAI 0.52%). This effect was not driven by any particular patient (**Supp. Fig. 5A**, **see Methods**). The PLC percentage was low (<0.1%), but the difference between sAI and non-sAI was highly significant and held both at the pre-treatment and post-treatment timepoint (**Fig. 2C**). Based on marker expression (**Fig. 2D-J**), PLCs strongly resembled platelets, but expressed two additional surface markers at high levels that are not ubiquitously associated with platelets: SPARC and TREML. Platelets are common contaminants in PBMC preparations^7^, however they are small and not expected to contain RNA. We hypothesized that this cell type constituted platelet lineage cells (PLCs) with special physical characteristics (e.g., larger size and transcript content) amenable to sc-RNAseq capture.

**Figure 2:**
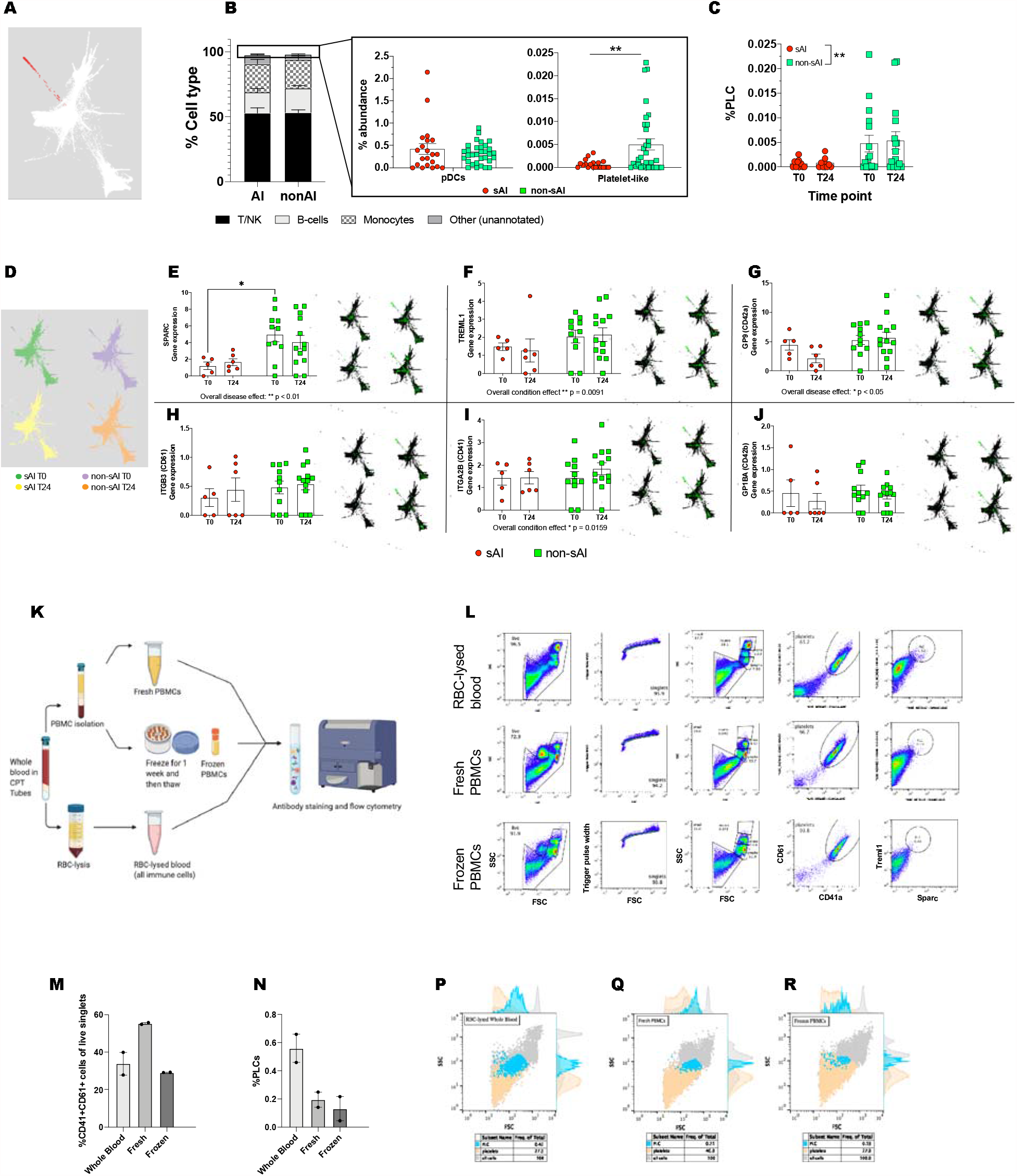
Platelet lineage cells (PLCs) are preferentially depleted in PBMCs of patients with secondary autoimmunity following alemtuzumab treatment. **A**. SPRING plot depicting single-cells of all samples in white, and unknown rare cell type in red. **B**. Bar graph depicting relative abundance of all major cell types as well rare cell-types: pDCs and platelet-like cells. Scatter bar plot depicting rare cell-type identified as Platelet-like cells is significantly enriched in non-sAI patients compared to sAI patients (Mean±S.E.M; Student’s t-test \*\**p*<0.01). **C**. Scatter bar plot showing that platelet-lineage cells (PLCs) are enriched in non-sAI patients at both timepoints: T0 and T24 (Mean±S.E.M; Two-way ANOVA, overall effect of AI state \*\**p*<0.01). **D**. SPRING plot depicting single cells separated and colored based on disease group (sAI T0: green, non-sAI T0: purple, sAI T24: yellow and non-sAI T24: orange). **E-J**. Scatter plots and accompanying SPRING plots showing the gene expression of SPARC, TREML1, GP9, ITGB3, ITGA2B, and GP1B, respectively (Mean±S.E.M; Two-way ANOVA with Sidak’s multiple comparison test \**p* < 0.05, \*\**p* < 0.001 and \*\*\**p* < 0.001). **K**. Flow cytometry workflow for investigating the frequency of PLCs in healthy donor fresh PBMCs, frozen PBMCs, and RBC-depleted whole blood. Samples were stained with CD41, CD61, Sparc, and Treml1 antibodies and analyzed on a BD Influx flow cytometry and cell sorting machine. **L**. Gating strategy to identify and quantify platelets and PLCs using FlowJo v10. **M, N**. Bar graphs depicting percentage of platelets and PLCs in whole blood, fresh PBMCs, and frozen PBMCs by flow cytometry (Mean±S.E.M). **P-R**. FSC-SSC Flow plots depicting size and granularity characteristics of all immune cells (grey), platelets (orange), and PLCs (blue).

To investigate their identity, we carried out a FACS experiment in samples from 2 healthy donors to estimate relative PLC abundance throughout sample isolation and processing steps (**Fig. 2K**). We designed a marker panel including the platelet markers CD41, CD61, and the specific markers SPARC and TREML1. We then applied this panel to whole blood, freshly isolated PBMCs, and frozen PBMCs from 2 healthy donors using the flow strategy described in **Fig. 2L**. All processing steps, except for storage duration, were done in accordance with the protocol used to preserve the sample set used in the clinical study. We observed that the proportion of CD41+CD61+ cells was 35% in whole blood, increased to 55% in fresh PBMCs (after removal of neutrophils), and decreased to 30% in frozen PBMCs (**Fig. 2M**). However, PLCs (defined as CD41+CD61+SPARC+TREML1+) constituted a mere 0.55% of whole blood and 0.1-0.2% of fresh and frozen PBMCs (**Fig. 2N**, **Supp. Fig. 5B**). To better understand the physical characteristics of PLCs in terms of size and granularity, we analyzed them with SSC/FSC gating, and showed they tended to be larger and more granular when compared to SPARC-TREML1-(double-negative) platelets (**Fig. 2P-R**). This is consistent with our hypothesis that these cells might appear in sc-RNAseq data due to their special physical characteristics.

Having elucidated the identity of this novel cell-type, we performed clustering of PLC transcriptomes to determine if there were any qualitative differences in PLCs prior to alemtuzumab treatment between patients with thyroid events (**Fig. 3A**). We found that there were distinct subsets of PLCs which differ in their expression of several markers and in their relative proportion (**Fig. 3B**). Subset 1 encompasses most PLCs in sAI patients, represented by 5 patients. It is characterized by lower expression of platelet markers, and higher expression of PDGFA, inhibitory markers (PDCD10), and nuclear proteins (DAB2, RGS10, RGS18, and TSC22D1). The second subset is relatively higher in the actin genes ACTB and ACTG1, and growth factor, and a potent chemoattractant and activator of neutrophils PPBP. This subset was also enriched in SPARC and TREML1 gene expression. These programs suggest a difference in maturity and activation state between the two subsets, with the subset depicting immaturity/resting state of PLCs being enriched in those with thyroid events (**Fig. 3C**). Therefore, we hypothesized that patients with or without thyroid events may differ in clinical measures of platelet maturity. To test this hypothesis, we carried out an analysis of Immature Platelet Fraction (IPF) data available for our cohort (Supp. File 1) and found that patients with thyroid events had a significantly increased IPF at T0 compared to those without (**Fig. 3D**). Moreover, patients with thyroid events tracked higher in IPF in monthly measurements taken throughout the two-year period (**Fig. 3E**). High IPF at T0 is not specific to sAI. However low relative PLCs and high IPF together appear to be a more specific indicator than high IPF alone (**Fig. 3F**, **Supp. Fig. 5C**).

**Figure 3:**
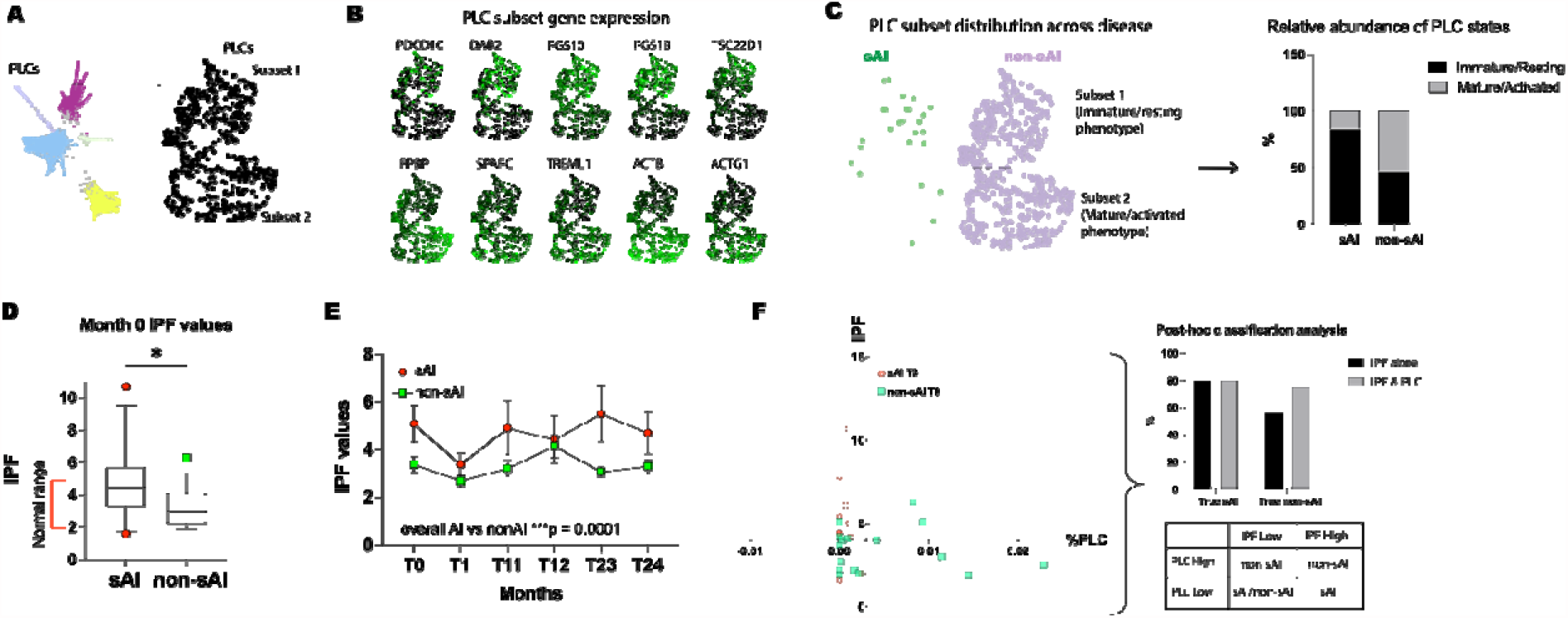
PLC subsets and correlation with immature PLC fraction in patients with secondary autoimmunity following alemtuzumab treatment. **A**. SPRING plot depicting selection and generation of subplot for PLCs from the main dataset. Subplot shows two clusters, Subset 1 and Subset 2, of PLCs separated based on transcriptomic differences. **B**. SPRING plots depicting expression of distinct genes enriched in Subset 1: PDCD10, DAB2, RGS10, RGS18, and TSC22D1; and in Subset 2: PPBP, SPARC, TREML1, ACTB, and ACTG1. **C**. Distribution of number of PLCs across the two subsets in sAI (green) and non-sAI (purple) patients, and bar chart depicting the % of cells within sAI and non-sAI groups that belong to Subset 1 (immature/resting transcriptomic state) or Subset 2 (mature/activated transcriptomic state). **D**. Box and whisker plot of immature platelet fraction (IPF) clinical values at month 0 in sAI and non-sAI patients, and normal range in red bracket (error bars span 10-90 percentile range). **E**. IPF clinical values over time in sAI and non-sAI patients (Mean±S.E.M.; Two-way ANOVA, overall sAI state difference *** p = 0.0001). **F**. Correlation graph of IPF clinical values and %PLCs from single-cell data for sAI and non-sAI patients at month 0. **Upper:** Post-hoc analysis of true sAI and true non-sAI identification of patients within the current cohort, based on either IPF values alone or IPF and %PLC values together. **Lower:** Tabular depiction of combinatorial use of clinical IPF values and single-cell PLC data in identifying AI status prior to treatment in the given cohort of patients.

To investigate how PLC deficit relates to subclinical manifestations of sAI, we generated data from three additional patients in the cohort who did not present with clinical sAI events but had detectable autoantibodies: two had anti-PLGLY antibodies, the other anti-TPO antibodies (see **Supp. File 1**). Interestingly, PLCs were absent in all three samples (**Supp. Fig. 5D**).

We also sought to examine whether the PLC numbers at T0 could be indicative of AI outcome beyond the 4-year horizon we originally set out to examine. Therefore, we obtained 7-year follow-up data for the same patients. We found that the 7-year data was largely consistent with the 4-year data with the exception of a single patient who developed AI sometime after the 4-year follow-up. Intriguingly, we observed this patient (10863747) had drastically lower PLCs at T24 compared to T0 (Supp. Fig. 5A).

Finally, we investigated whether platelet and PLC-specific gene expression can distinguish between AI and non-AI patients outside of the small set of samples we obtained for scRNA-seq. To this end, we queried a bulk RNA-seq dataset previously generated from PBMC samples of 161 MS patients from two cohorts treated with alemtuzumab as part of the same trial at three timepoints (baseline, month 12 and month 24, **Fig 4a**). The patients in this dataset were roughly evenly split between AI(76), and non-AI(75). We defined a general set of PLC/platelet-relevant genes by combining known platelet markers (**Fig. 2h-j**) and PLC subset-specific genes (**Fig 3b**, excluding ubiquitously expressed Actin B and G). When we co-clustered the baseline bulk RNA-seq expression profiles from the patients with this general gene set, we obtained two top-level patient clusters (Cluster A and Cluster B) encompassing 160 out of the 161 samples (**Fig 4b**). Cluster A was defined by the relative downregulation of mature PLC/patelet markers, which formed their own gene cluster. We observed that this cluster was predominantly (58%) made up of AI patients, while Cluster B (marked with a black tripe) was predominantly (60%) non-AI (*p < 0.04 by Fisher exact test, OR > 2). Thus, the risk of a secondary autoimmunity adverse event for patients in Cluster A appeared doubled. Similarly, when we analysed the gene expression of the mature PLC/platelet marker cluster in AI patients, we noticed it was downregulated across all three timepoints (ANOVA F-test (Sampling Time + AIvsNonAI); **p < 0.004 AI vs non-AI; sampling time and interaction n.s., sampling time p = 0.31; interaction p = 0.53), while the other “immature” PLC genes appeared to be only marginally downregulated(ANOVA F-test (Sampling Time + AIvsNonAI); AI vs non-AI n.s. (p = 0.0999); sampling time and interaction n.s., sampling time p = 0.70; interaction p = 0.42). This was consistent with our previous observation that high immature platelet fraction is associated with AI.

**Figure 4:**
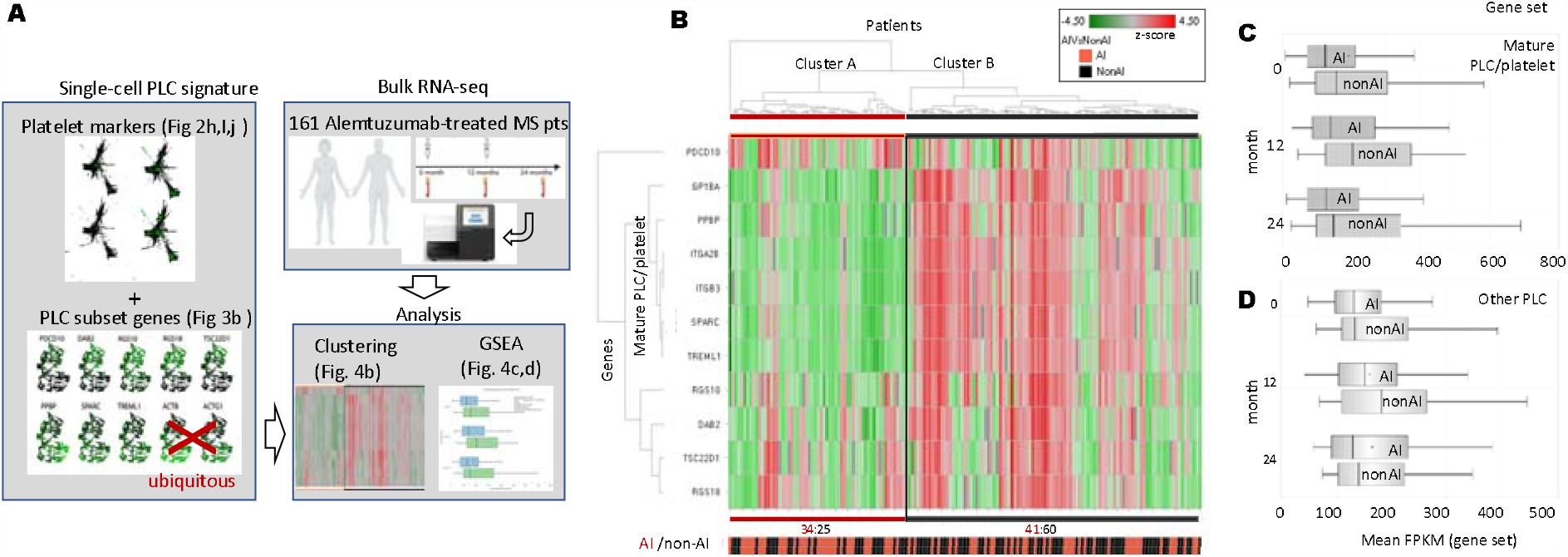
PLC signature expression analysis of bulk RNA-seq data from Alemtuzumab-treated cohort. **A**. Workflow of figure 4: Platelet markers and PLC subset genes (excluding ubiquitously expressed Actin B and G) were used to define a PLC signature; co-clustering of bulk RNA-seq expression profiles from 161 Alemtuzumab-treated patients with PLC signature and a gene set enrichment analysis across patient groups was performed. **B**. Clustering analysis of the PLC signature in bulk RNA-seq expression profiles from 161 Alemtuzumab-treated patients (76 AI, 75 non-AI) at baseline. Gene expression (top) is shown as z-score across all patients; AI status (bottom) is indicated in red (AI) and black (non-AI). Cluster A and Cluster B emerge as top-level clusters encompassing 160 out of 161 total samples. Cluster A (marked by a red stripe) is defined by the relative downregulation of mature PLC/patelet markers and is predominantly (58%) made up of AI patients, while Cluster B (marked with a black tripe) is predominantly (60%) non-AI (*p < 0.04 by Fisher exact test). **C**. Mean gene expression of mature PLC/platelet markers downregulated in cluster A across all three timepoints in AI and non-AI patients (ANOVA F-test (Sampling Time + AIvsNonAI); **p < 0.004 AI vs non-AI; sampling time and interaction n.s. (*sampling time p* = 0.31; *interaction* p = 0.53). **D**. Mean gene expression of non-mature PLC/platelet markers downregulated in cluster A across all three timepoints in AI and non-AI patients (ANOVA F-test (Sampling Time + AIvsNonAI); AI vs non-AI n.s. (p = 0.0999); sampling time and interaction n.s. (*sampling time* p = 0.70; *interaction* p = 0.42).

## Discussion

The efficacy of alemtuzumab for the treatment of RMS has been established in several clinical trials^8^. In many cases, patients who have failed other treatments have achieved lasting benefits of up to 7 years post follow up^9^. However, the common occurrence of secondary thyroid events in alemtuzumab-treated patients observed in the clinical trials, presents a concern. Furthermore, 2.2% of patients presented with ITP in the clinical development program with 0.4% being reported in real-world clinical practice^2^. Despite extensive investigation prior to this study^10^, there were no identifiable patient characteristics at the start of treatment that could be linked to secondary autoimmunity.

Using high-resolution droplet-based scRNA-seq, we discovered that a hitherto uncharacterized subset of platelet lineage cells (PLCs) is negatively associated with secondary thyroid autoimmunity. We also observed a difference in maturity and activation state in PLCs of sAI and non-sAI patients, which we linked to a measurable clinical parameter – IPF, and validated our findings on a larger set of patients using a co-clustering analysis of samples with the PLC signature defined from single-cell data. While scRNA-seq is far from becoming a scalable clinical assay, and IPF is not a sufficiently specific indicator of sAI, our findings suggest that developing clinical grade assays focused on PLCs may prove useful. They also raise the interesting possibility that pre-existing subclinical platelet deficiency is linked to secondary autoimmunity following alemtuzumab treatment either directly, or through a common underlying mechanism.

To perform this study, we overcame logistical, technical, and computational challenges^11^. Understandably, the samples collected nearly a decade ago were neither prepared nor banked with downstream single-cell applications in mind, and the majority had already been allocated to other analytic techniques. Unfortunately, the samples remaining did not include any patients who developed more severe secondary autoimmunities such as ITP, although our analysis of three patients with lab abnormalities suggests an anecdotal link to the presence of autoantibodies against platelets as well as thyroid. Nevertheless, this study demonstrates the broad potential of scRNA-seq to enable biomarker discovery and generate novel insights where standard methods have failed to yield results.

## Data Availability

Data will be available upon final publication

## Acknowledgements

We would like to thank Dr. Alexei Protopopov, Dr. Emma Wang, and Maximilian Rogers-Grazado in Precision Oncology, Sanofi, Cambridge, MA, for their help in performing the sequencing of libraries. We would also like to thank Dr. Angelique Biancotto for supervising the flow cytometry experiments.

---

**Supplementary Figure 1.**
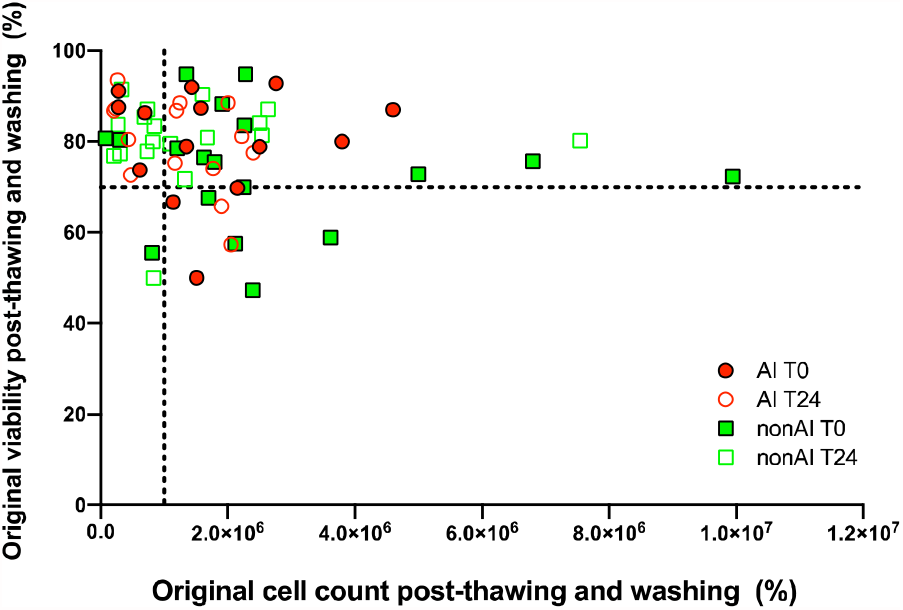
Cell count and viability of patient PBMCs post-thawing. PBMCs were thawed and counted using a hemacytometer. X-Y Correlation graph shows a wide range of cell counts and viability of PBMC samples post-thaw for every sample. Samples with viability lower than 75% were processed for “dead cell removal” (see **Methods**).

**Supplementary Figure 2.**
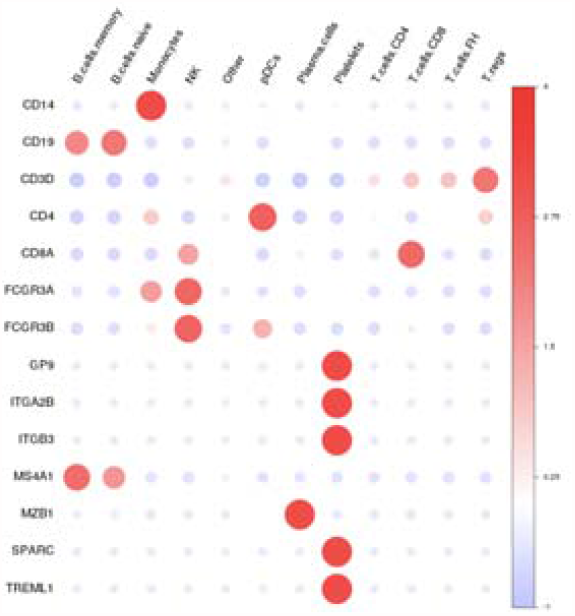
Gene expression of typical markers of identification for different cell types. Heatmap depicts the relative expression of typical gene markers expressed on cell-types annotated by the automatic cell-type identification tool within the single-cell dataset. Color scale shows average expression of each gene normalized across cell types.

**Supplementary Figure 3.**
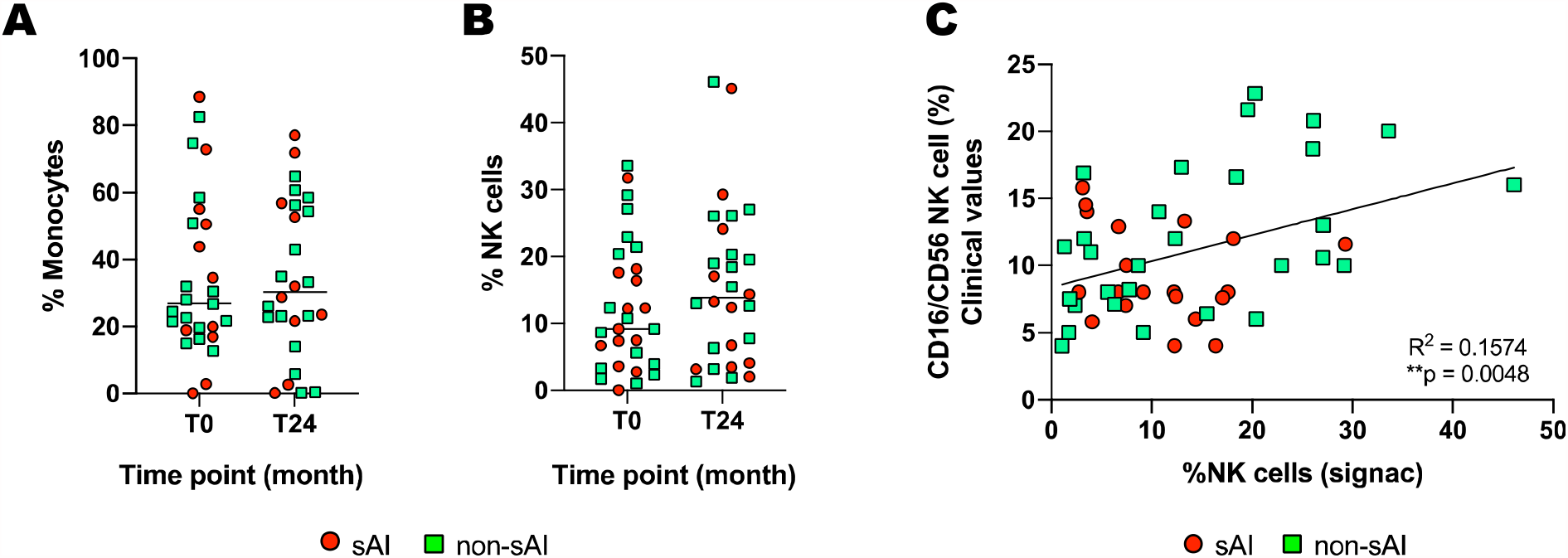
Alemtuzumab treatment does not affect the number of monocytes or NK cells in sAI or non-sAI patients. **A, B**. Scatter bar plot shows single-cell data of % of monocytes and % of NK cells before and after alemtuzumab treatment. C. Correlation graph of clinical values (flow cytometry) and single-cell numbers % NK cell numbers for all patients. Pearson’s correlation R^2^=0.212, *p=0.0025.

**Supplementary Figure 4.**
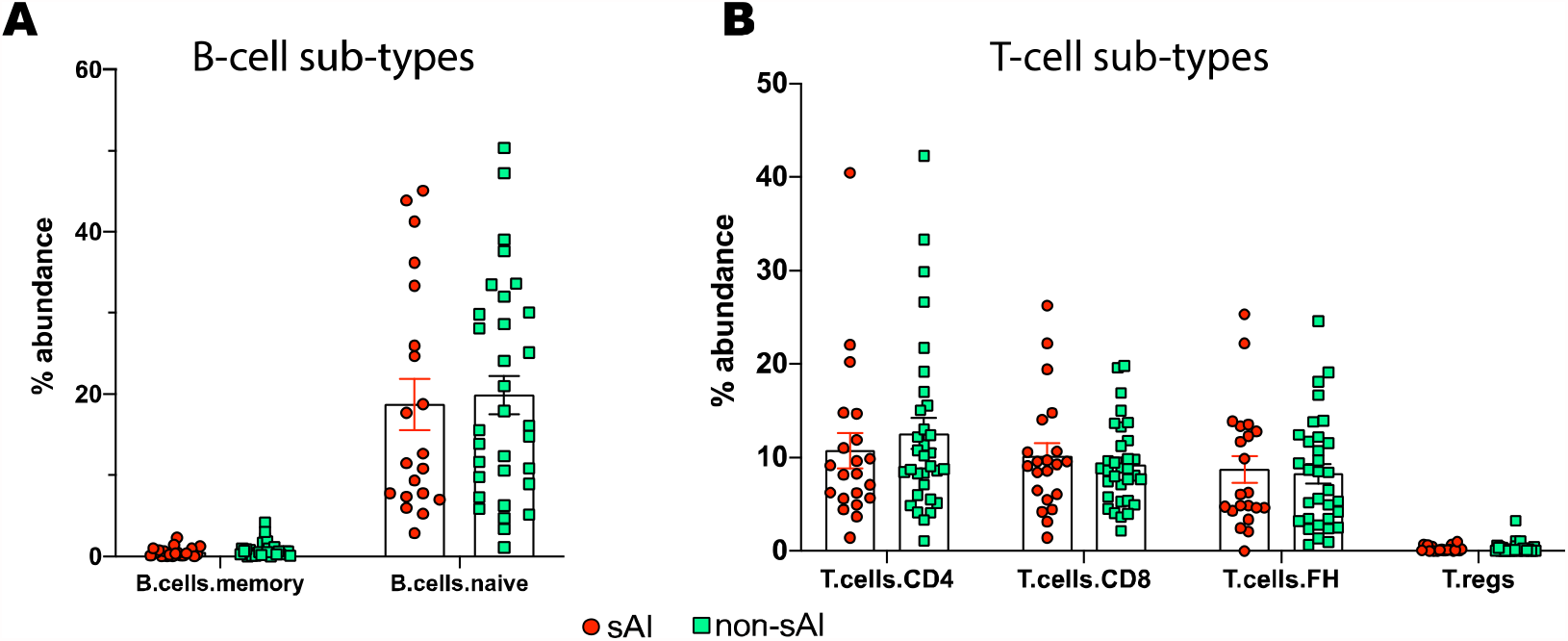
There is no difference in number of B and T cell subsets between sAI and non-sAI patients. **A**. Scatter bar plot depicting single-cell data of % naïve and memory B cells and **B**. % CD4 T cells, CD8 T cells, T follicular helper cells (TFH), and Regulatory T cells (T-regs) in sAI and non-sAI patients (Mean±S.E.M).

**Supplementary Figure 5.**
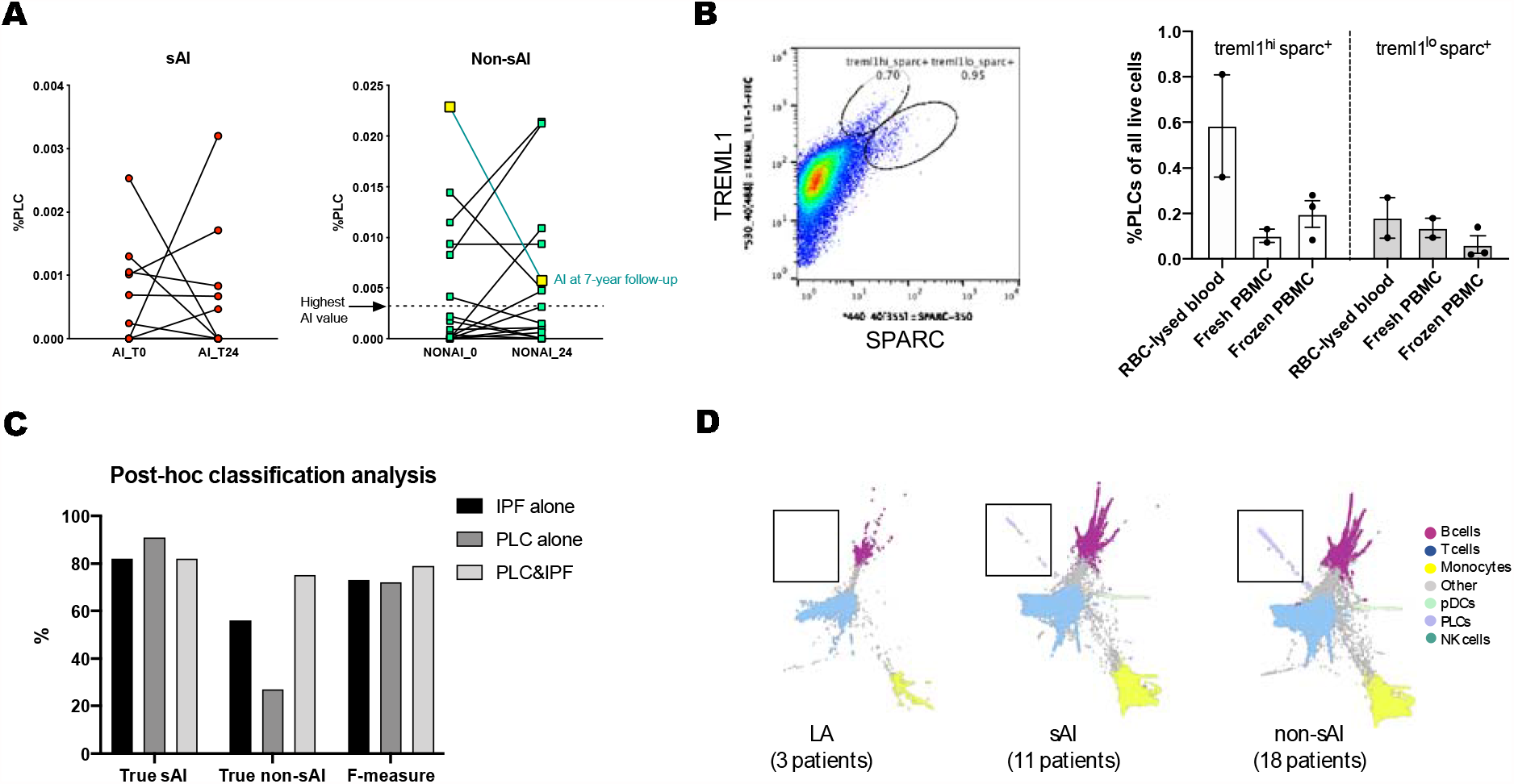
Characterizing PLC phenotype in alemtuzumab treated patients. **A**. Scatter plot depicting individual patient values of %PLCs from sc-RNAseq data before and after alemtuzumab treatment in sAI and non-sAI patients. The non-sAI patient highlighted in yellow showed reduced PLC numbers at T24, and developed sAI between 4 and 7 years of follow up. **B**. Representative flow cytometry plot of Sparc and Treml1 positive platelets in the RBC-lysed blood shows two lobes: 1. Treml1^hi^ sparc^+^ and 2. Treml1^lo^ and sparc^+^. Bar plot quantifying the % of PLCs in the two lobes separately in RBC-lysed blood, fresh PBMCs, and frozen PBMCs. **C**. Post-hoc analysis of patients in the current study showing performance of IPF values alone, PLC alone, or IPF and PLC together in identifying true sAI or true non-sAI status. **D**. SPRING plot of single-cell data of patients with either laboratory abnormalities (LA), secondary autoimmunity (sAI), or no secondary autoimmunity (non-sAI).

## Methods

### Clinical trial and sample collection

Cryopreserved PBMC samples were obtained from the CAMM323 study (<u>CARE-MS I</u>, Clinicaltrials.gov identifier NCT00530348) with full informed consent. In this study, patients diagnosed with relapsing remitting multiple sclerosis (RR-MS) and who had not been previously treated with an MS disease-modifying therapy were treated with alemtuzumab (12 mg/day, IV) for 5 days at baseline (T0) and for 3 days 12 months later (T12), or with subcutaneous interferon beta-1a 44 µg 3× weekly. Clinical metadata for the patients is presented in supplementary file 1. Whole blood (6-8 mL) was collected in CPT tubes with sodium citrate at T0, T12 (12 months post first course), and T24 timepoints. This study analyzed samples from T0 and T24 time points for a total of 32 patients of which 11 developed secondary autoimmunity (sAI, defined by occurrence of thyroid events) following alemtuzumab treatment, 18 who did not (non-sAI), and 3 patients who had laboratory abnormalities (LA) as defined by the presence of autoantibodies^1^. In the adjudication, a thyroid event was defined as either having a laboratory finding (e.g. abnormal TSH) or a clinical adverse event (AE). If diagnosed, the clinical AE was classified as either Grave’s disease (i.e. hyperthyroidism), Hashimoto’s disease (i.e. hypothyroidism), transient thyroiditis, Grave’s disease switching to hypothyroidism, or Hashimoto’s disease switching to hyperthyroidism, or uncertain. No demographic co-variate such as age, sex or BMI showed associations with sAI (data not shown). The timeline for autoimmunity development in described in previous studies^2,3^.

### PBMC processing and storage

Following blood collection, the CPT tube was centrifuged at the clinical site, such that the RBCs were captured within the gel barrier. The plasma layer and white buffy coat of PBMCs was mixed together prior to shipment. CPT tubes were shipped to the laboratory at room temperature and processed within 60 hours of collection. PBMC collection was carried out in a class II biological safety cabinet. Cells were inverted into the plasma gently 5-10 times. CPT tubes were opened and the entire suspension above the gel was transferred into a sterile 15 mL conical tube. The volume of the solution was noted. Samples were centrifuged at 300 rcf for 10-15 minutes. Plasma was removed and discarded without disturbing the cell pellet. The pellet was resuspended by gentle pipetting. Dulbecco’s PBS (1X) was added to make up the volume to 10-13 mL. Samples were then centrifuged for 10-15 min at 300 rcf. The supernatant was aspirated without disturbing the pellet. Dulbecco’s PBS (1X) was added to bring volume to 10 mL. Samples were inverted and mixed gently. White blood cell count, and % lymphocyte and % monocyte counts were determined using a Gen-S hematology analyzer. Cell viability was determined by staining with Propidium Iodide and processing on the FACSCalibur™. Following this, samples were centrifuged for 10-15 min at 300 rcf and the supernatant was aspirated. The pellet was gently resuspended by pipetting. 2 mL of freezing solution Cryostor CS10 (Stemcell Technologies, Cat # 07930) was added and mixed using a 1 mL pipet. 1 mL of cell solution was aliquoted into 2 cryovials and stored at -80°C for a minimum of 24 hours and a maximum of 72 hours before long-term storage in liquid nitrogen.

### PBMC thawing and pre-processing for single-cell workflow

As described previously in our single-cell optimization study^4^, cryopreserved PBMCs were thawed (2 vials at a time) in a 37°C water bath for 1-2 minutes until a small crystal remained. The cryovial was removed from the water bath and cell solution was transferred to a sterile 2 mL Eppendorf tube using a wide bore pipet tip. The cryovial was washed with 1 mL of 0.04% BSA/PBS and the solution was transferred to the Eppendorf tube. The sample was centrifuged at 150 rcf, 5 min, at room temperature (RT). The supernatant was carefully removed, and the sample was washed with 1 mL of 0.04% BSA/PBS using wide bore pipet tip. The sample was washed twice more using the same conditions above, for a total of 3 washes. After the final wash, cells were resuspended in 1 mL of 0.04% BSA/PBS and counted using a hemacytometer (C-Chip, SKC, Cat # DHCF015) with trypan blue (0.4%, GIBCO Cat #15250061) as the stain. If the viability was found to be lower than 75%, the sample was subjected to a “clean-up” step using Dead Cell Removal kit (Miltenyi Biotec, Catalog #130-090-101). Cells were washed again and resuspended in 500 µL of 0.04% BSA/PBS and counted. The volume was adjusted to 1 × 10^6^ cells/mL of 0.04% BSA/PBS.

### Single-cell transcriptomics

After the cell volume was adjusted to 1 × 10^6^ cells/mL, the protocol for 10X Genomics 5’ gene expression library preparation was used. 8000 cells were targeted per sample. Quality of uniquely-indexed libraries was determined using the 2100 Bioanalyzer instrument (Agilent) with a High Sensitivity DNA kit (Agilent, Catalog # 5067–4626) and quantified using Kapa library quantification kit (Kapa Biosystems, Catalog # KK4824 – 07960140001) on the QuantStudio 7 Flex Real-Time PCR system. The libraries were diluted in 10 mM Tris-HCl buffer (pH 8.0) and pooled in equimolar concentration (2 nM) for sequencing.

Sequencing was carrid out on a NOVAseq6000 system (Illumina), using NOVAseq 6000 S2 reagent kit (300 cycles, Illumina Cat #20028314). Sequencing depth and cycle number was as per 10X Genomics recommendations: Read 1⍰=⍰26 cycles, i7 index⍰=⍰8 cycles, Read 2⍰=⍰98 cycles, and we aimed for a sequencing depth of 35,000 reads per cell.

### Single-cell pipeline, preprocessing and QC

Single cell analysis was carried out as described previously^4^. Briefly, following BCL conversion, FASTqs were processed through CellRanger version v2.1.1 for demultiplexing, alignment, filtering, barcode counting, UMI counting, and generating of gene x barcode matrices. Following this, an in-house built pre-processing pipeline was run for background removal of ambient RNA and filtering out empty barcodes as well as stress and mitochondrial genes. The resulting hdf5 files were then input into an internal single-cell visualization tool called SPRING^5^. SPRING also allows for compressing/reducing the size of large single-cell datasets for fast visualization. An in-house developed tool was then used to automatically annotate cell-types and cell-subtypes, as well as can identify novel cell-types. After close scrutiny, bad quality samples were excluded from analysis. Briefly, CellRanger web summaries for each of the samples was evaluated. If the “cliff graph” was found to pass QC, sequencing depth was evaluated. For samples with low sequencing depth, the libraries were re-pooled to account for low-read samples and re-sequenced. Data from all sequencing runs was combined and re-run through CellRanger. Web summaries were evaluated again and samples with low cell counts and bad “cliff graphs” were flagged. All samples were processed through the internal pipeline for decontamination and filtering (as described above) and visualized on the SPRING portal. Samples that clustered poorly on the SPRING layout or had fewer than 100 cells were flagged. Thus, samples that failed CellRanger QC and SPRING clustering were eliminated from analysis. Seven such samples were identified and eliminated. The resulting new dataset was labeled Lemtrada_SC (sample clean-up). As these samples belonged to different timepoints of different patients, another dataset was created which eliminated paired samples from patients in Lemtrada_SC dataset. This new dataset was called as Lemtrada_PC (patient-clean-up). The information on sequencing reads, CellRanger QC, cell numbers per sample as well as SC & PC dataset details are described in Supplementary File 1.

### Bulk Transcriptomics

Bulk transcriptomic data from PBMC samples obtained in studies CAMM323 and CAMM32400507 was previously obtained using NGS Illumina Platform HiSeq2000 and standard PolyA library preparation methods. Data processing and analysis was performed within the OmicSoft Studio software (QIAGEN®, Redwood City, CA) as follows: RNA-seq data alignment was performed using OSA aligner to human genome version GRChg37. Quantification was performed using UTR-trimmed RSEM. FPKM values are first upper quartile normalized at the gene level, not counting mitochondrial genes or genes with an exon length < 500. The upper quartile target value is set to 10. Then FPKM values are log2 transformed by log2(FPKM+0.1). Only samples with RIN > 7.5 were considered for this analysis. HierarchicalClustering method used the default complete linkage, and Pearson correlation distance metric (default). No additional parameterization was explored.

### Flow Cytometry

#### Pre-processing of cells

Whole blood samples were obtained from two healthy subjects using Sanofi’s internal Donor Research Program in Framingham, MA. 75-100 mL of blood was collected per donor into CPT tubes with Sodium Citrate (BD Biosciences, Cat #362761). In the first experiment, PBMCs were isolated from the blood as described above in the “Clinical trial and sample collection” section. Following this, PBMCs were washed twice with 1× PBS and counted using propidium Iodide/Acridine Orange stain (Nexcelom, Cat # CS2-0106-5ML) on the Cellaca MX (Nexcelom, model MX-SYS1). Half of the freshly collected PBMCs were stored in Cryostor CS10 (Stemcell technologies Cat # 07930) and frozen at –80°C for 24 hours followed by 1 week storage in liquid nitrogen. After 1 week, cells were thawed, counted, and processed for flow cytometry. The remaining half of fresh PBMCs were processed for antibody staining and flow analysis. In the second set of experiments, the healthy donors were recalled, and blood was collected as above. RBC lysis was done using ACK lysis buffer (Gibco, A10492-01, lot# 2048611) as per manufacturer’s instructions. Briefly, two 50 mL Falcon conical tubes were used for each donor. Approximately 10-12 mL of whole blood was poured into each 50 mL tube after initial centrifugation to remove plasma. ACK buffer was added to 45 mL and incubations were on a VWR variable speed rocker for 10 minutes. After a 3^rd^ 10-minute incubation, the cell pellets were mostly white, indicating red cells had been lysed and removed by washing. After this, cells were processed for staining and flow cytometry.

#### Compensation, staining, and flow analysis

Before running samples on the BD Influx (Becton Dickinson Influx Configurable, model 646500, s/n X64650000137), auto-compensation was done according to the Influx protocol. Briefly, two drops of compensation beads (see below) and one test of stain were mixed, incubated for 20 minutes on ice in the dark and then resuspended in 350 µL stain buffer in 5 mL Falcon tubes. This was used to perform compensation on the BD Influx prior to running the samples.

All cells were centrifuged at 300g, 5 min, 4°C and pellets were resuspended in 3 mL BD stain buffer and transferred to a 5 mL Falcon polypropylene tube. Cells were centrifuged again and the pellet was resuspended in 100 µL of staining panel, and incubated on ice in the dark for 30 min. The staining panel consisted of: CD61-BV510 (5 µL/test), CD41A-APC (20 µL/test), TREML/TLT-1-FITC (5 µL/test), SPARC-(350) (5 µL/test). After incubation, 3 mL of stain buffer was added and centrifuged. The supernatant was removed and pellets were resuspended in 2 mL of stain buffer and analyzed on the BD Influx as per the manufacturer’s instructions. BD Influx information: Amplitude was set at 4.91, Drop Frequency was 44.70, stream focus was 15, Drop position was 200, Max drop was 101, Drop Delay was 28.43, and stream deflections for tubes was -84, -33, 33, 86.

| Name | Supplier | Catalog Number | Lot Number |
| --- | --- | --- | --- |
| Stain Buffer | Becton Dickerson | 554656 | 8145755 |
| Compensation Beads | Invitrogen | 01-2222-42 | 2037700 |
| Sphero ultra-rainbow fluorescent particles | Spherotech | URFP-30-2 | AK02 |
| FACS Accudrop beads | Becton Dickerson | 345249 | 822690 |
| PBS | Gibco | 14190 | 2004029 |
| CD41A-APC | Becton Dickerson | 559777 | 8332624 |
| CD61-BV510 | Becton Dickerson | 563303 | 9126835 |
| TREML/TLT-1-FITC | R&D Systems | FAB2394G | AFEQ0118111 |
| SPARC (350) | R&D Systems | IC941U-100ug | 15546582 |

#### Gating Strategy

Live cells were gated from the log scale on FSC and SSC, excluding dead cells. Of the live cells, “small cells” were gated using log scale on FSC and SSC because of their small size, 2-3 µm. Singlets were gated off the FSC and SSC gate using Trigger-pulse width. CD41A+, CD61+ were considered markers for platelets and were gated off the singlet gate. Off of the CD41A+, CD61+ gate, PLCs were identified by being double-positive for SPARC and TREML1.

### Statistical Analysis

FlowJo (version 10) was used for analyzing flow data. GraphPad Prism (version 8) was used for generating graphs and performing statistical analyses. Levels of significance are indicated by: ***p⍰<⍰0.00⍰, **p⍰<⍰0.0⍰, and *p⍰<⍰0.05.

PLC abundance results were tested for robustness against the presence of anti-PLGY antibody by excluding the single sAI patient (10553163) who also presented with this lab abnormality. Significance level was p<0.015 with this patient excluded.

We also observed that SPARC+TREML1+ cells showed bimodal distribution, with some cells showing higher TREML1 expression than others (**Supplementary Fig. 5B**). On quantifying the lobes separately, we observed no significant difference in relative distribution of these cells across the 3 different sample types.

